# Genome-wide analysis identifies significant contribution of brain-expressed genes in chronic, but not acute, back pain

**DOI:** 10.1101/2020.09.04.20187575

**Authors:** Andrey V. Bortsov, Marc Parisien, Samar Khoury, Dmitri V. Zaykin, Amy E Martinsen, Marie Udnesseter Lie, Ingrid Heuch, HUNT All-In Pain, Kristian Hveem, John-Anker Zwart, Bendik S Winsvold, Luda Diatchenko

## Abstract

Back pain is the leading cause of disability worldwide. Although most cases of back pain are acute, 20% of people with acute back pain go on to experience chronic back pain symptoms. It is unclear if acute and chronic pain states have similar or distinct underlying genetic mechanisms. Here we performed a genome-wide analysis for acute and chronic back pain in 375,158 individuals and found a significant genetic contribution to chronic, but not to acute, back pain. Using the UK Biobank cohort for discovery and the HUNT cohort for replication, we identified 7 loci for chronic back pain, of which 3 are novel. Pathway analyses, tissue-specific heritability enrichment analyses, epigenetic characterization, and tissue-specific transcriptome mapping in mouse pain models suggest a substantial genetic contribution to chronic, but not acute, back pain from the loci predominantly expressed in the central nervous system. Our findings show that chronic back pain is more heritable than acute back pain and is driven mostly by genes expressed in the central nervous system.

## INTRODUCTION

Back pain is the world’s leading cause of disability, reducing quality of life, and imposing significant health care costs.^1–4^ According to the International Association for the Study of Pain (IASP) criteria, a back pain episode lasting less than 3 months is defined as acute back pain while pain lasting for 3 months or longer is defined as chronic.^5^ Although the majority of individuals with acute back pain resolve their pain,^6,7^ a substantial proportion of these cases transition to chronic back pain. As a result, chronic back pain is the most common painful condition which affects 10–15% of the adult population. ^2,3,8^ Mechanisms involved in the development of chronic back pain remain largely unknown, and seem to be different from causes leading to the onset of acute back pain. While some believe that the majority (up to 85%) of cases of chronic back pain cannot be attributed to a known specific pathology,^9^ others attribute chronic back pain to processes such as central sensitization,^10^ disc herniation causing inflammation of the affected nerve,^11^ facet joint syndrome,^12^ sacroiliac joint pain,^13–15^ lumbar spinal stenosis^16^, and lumbar disc degeneration.^17^

Heritability estimates for back pain from twin studies range from 30 to 68%.^18–21^ Twin studies also show shared heritability between chronic back pain and lumbar disc degeneration,^18^ depression,^22^ anxiety,^22^ education,^23^ obesity,^24^ chronic widespread musculoskeletal pain.^25^ Recent genome-wide association studies (GWAS) using the UK Biobank cohort have identified a number of genes associated with chronic back pain, including *SPOCK2, SOX5*, and *DCC*.^26,27^ These studies confirm genetic involvement both in the etiology of chronic back pain and its related comorbidities.

Despite the mounting evidence that acute and chronic back pain are distinct states with different pathologies, there is a significant gap in knowledge about the genetic risk factors and molecular pathophysiology associated with acute versus chronic back pain. Previous genetic studies have looked either at back pain in general,^26^ or only at the chronic pain condition.^27^ One important question is whether acute and chronic pain states have similar underlying genetic pathways. Here we used GWAS for both acute and chronic back pain states and characterized the corresponding molecular and cellular pathways contributing to these genetics assotiations. We hypothesized that acute and chronic back pain were different with respect to individual genetic risk loci, overall genetic heritability, heritability distribution across tissues and cells, and contributing molecular and cellular pathways.

## RESULTS

In our genome-wide exploration of genetic factors contributing to acute versus chronic back pain we used data on individuals of white British ancestry from the UK Biobank cohort (n=375,158) (Fig. 1). Acute and chronic back pain phenotypes were inferred from an affirmative response to the question of whether the respondent had experienced back pain in the last month that interfered with their usual activities. Those who reported back pain in the last month were asked if their back pain lasted for more than 3 months. Acute back pain was defined as back pain (that interfered with activities) in the last month that lasted no more than three months. Chronic back pain was defined as back pain that lasted for more than three months. Individuals with no acute or chronic back pain (but possibly with other types of pain) were used as controls. The defined phenotypes were used as outcome variables to perform the GWAS.

**Figure 1.**
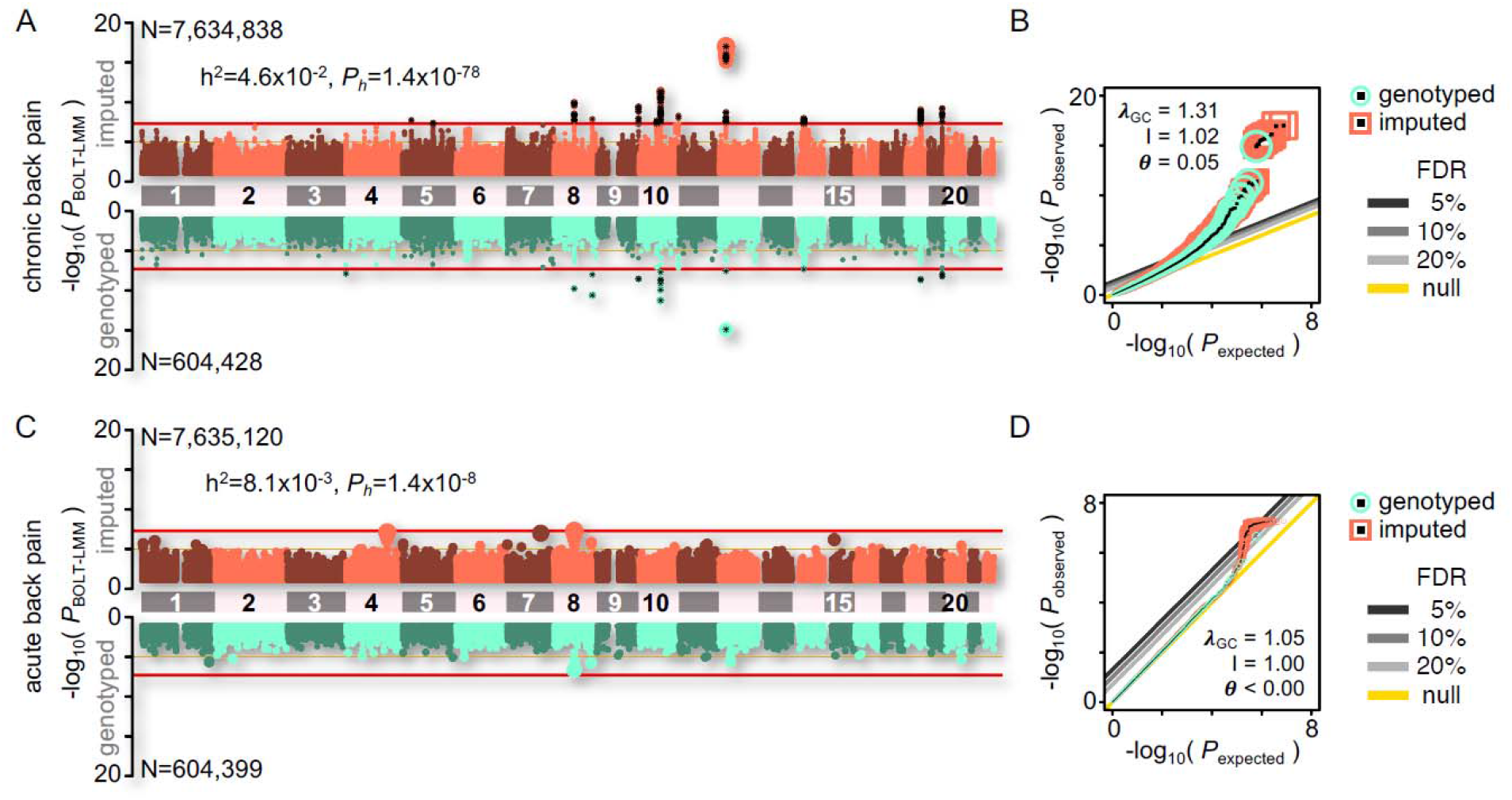
Genome-wide association analysis of chronic back pain (A, B) and acute back pain (C, D) in 375,158 participants from the UK Biobank cohort. **(A, C)** Miami plots show –log10 P-values for imputed (top, orange) and genotyped (bottom, green) SNPs (N, number of analyzed subjects). Horizontal red lines indicate genome-wide statistical significance at 5×10^−8^. Genetic heritability estimate (h^2^) shown with corresponding P-value (*P*_h_). **(B, D)** QQ plots show observed –log10 P-values (*P*_observed_) as a function of expected –log10 P-values (*P*_expected_), for both genotyped (green) and imputed (orange) SNPs. Genomic control parameter (*λ*_GC_), LD Score regression intercept (I) and their ratios (*θ*) are shown.

A genome-wide association analysis of chronic back pain (n=375,158; 70,633 cases; 304,525 controls) revealed 355 SNPs that reached the genome-wide significance threshold (5×10^−8^), grouped into 13 loci (Fig. 1A, Supplementary Table 1). The majority of the significant loci (8 of 13) have already been reported as significantly associated with back pain in the UK Biobank cohort by other groups (chr4: TMED11P • SPON2, chr8: C8orf34-AS1 • C8orf34 • LINC01592, chr8: CCDC26 • GSDMC, chr9: NRARP • EXD3 • NOXA1, chr10: PSAP • CHST3 • SPOCK2 • ASCC1, chr10: BNIP3 • JAKMIP3 • DPYSL4, chr12: LOC101928441 • SOX5 • SOX5-AS1, chr18: LINC01630 • DCC • LINC01919), and five loci were new (chr5: LOC105374704 • CDH6, chr5: NUDT12 • RAB9BP1, chr10: LINC00844 • CCEPR, chr14: SLC25A21-AS1 • MIPOL1 • FOXA1, chr19: BCAM • NECTIN2 • TOMM40 • APOE • APOC1 • APOC1P1).^26,27^ The estimated narrow-sense SNP heritability (h^2^) for chronic back pain was 4.6% (*P*=1.4×10^−78^, Fig. 1A). While there was some inflation of test statistics (genomic control parameter λ_GC_ = 1.31), the LD Score regression intercept was close to 1 (Fig. 1B), indicating that the inflation was due to polygenicity rather than confounding factors such as population stratification and/or cryptic relatedness, the use of a mixed model, or population stratification.^28^ The latter confounding factors were mitigated using BOLT-LMM via genetic principal components and kinship covariance matrices.^29,30^

The GWAS of acute back pain (n=336,734; 32,209 cases and 304,525 controls) revealed no genetic variants reached the genome-wide statistical significance threshold (Fig. 1C). The estimated narrow-sense SNP heritability (h^2^=0.81%) for acute back pain was also lower than for chronic back pain; however, the heritability was significantly different from zero (*P*=1.4×10^−8^, Fig. 1C). The LD Score regression intercept was 1, suggesting that the moderate inflation (λ_GC_1.05) was due to polygenicity rather than confounding factors (Fig. 1D). Of note, amongst the 13 statistically significant loci in chronic back pain, only one locus on chromosome 8 was associated with both acute and chronic back pain (Supplementary Table 2).

In the HUNT replication cohort, we found similar differences in heritability estimates between acute and chronic back pain. Heritability of chronic back pain was 3.4% (*P*=0.0011), whereas heritability of acute back pain was 0.6%(*P*= 0.851). From this we concluded that the heritability of acute back pain is approximately 5–6 times smaller than the heritability of chronic back pain. A total of 7 out of 13 statistically significant loci were replicated in the HUNT cohort for chronic pain cases (Table 1) at the P < 0.05 level using sets of SNPs at high LD (r > 0.8).

**Table 1.** SNP-level discovery and replication results of chronic back pain GWAS. Replication of the top hit within each locus was performed using SNP sets in high LD (r2 > 0.8). SNPs with lowest replication P-values for each locus in the discovery and replication cohort are shown.

|  |  |  | Discovery, top hit |  |  | Replication |  |  |  |  |
| --- | --- | --- | --- | --- | --- | --- | --- | --- | --- | --- |
|  | Chr | Genes Cluster | SNP | BP | P | SNP | BP | P | r2 | Direction |
| 1 | 4 | LOC105374344•TMED11P•SPON2 | rs6826705 | 1122634 | 1.30E-08 | rs4368552 | 1128054 | 0.067 | 0.97 | ↑↑ |
| 2 | 5 | LOC105374704•CDH6 | rs2066928 | 30843787 | 1.80E-08 | rs2066928 | 30843787 | 0.034 | 1 | ↑↑ |
| 3 | 5 | NUDT12•RAB9BP1 | rs325485 | 103995368 | 3.90E-08 | rs325528 | 104048590 | 0.0065 | 0.88 | ↑↑ |
| 4 | 8 | C8orf34-AS1•C8orf34•LINC01592 | rs1865442 | 69574165 | 1.10E-10 | rs72666774 | 69587018 | 0.0042 | 0.96 | ↑↑ |
| 5 | 8 | CCDC26•GSDMC | rs12056383 | 130711839 | 1.00E-08 | rs77753889 | 130711599 | 0.631 | 0.85 | ↑↓ |
| 6 | 9 | NRARP•EXD3•NOXA1 | rs28458909 | 140257189 | 2.90E-09 | -- | -- | -- | -- | -- |
| 7 | 10 | LINC00844•CCEPR | rs12415581 | 60899641 | 2.10E-08 | rs7084967 | 60945358 | 0.356 | 0.80 | ↑↑ |
| 8 | 10 | PSAP•CHST3•SPOCK2•ASCC1 | rs1668172 | 73823019 | 4.10E-12 | rs1245561 | 73837350 | 0.0046 | 0.98 | ↑↑ |
| 9 | 10 | BNIP3•JAKMIP3•DPYSL4 | rs10870267 | 133968063 | 5.00E-09 | rs34588848 | 134005567 | 0.480 | 0.83 | ↑↑ |
| 10 | 12 | LOC101928441•SOX5•SOX5-AS1 | rs12310519 | 23975219 | 9.50E-18 | rs56290807 | 23972014 | 0.035 | 0.97 | ↑↑ |
| 11 | 14 | SLC25A21-AS1•MIPOL1•FOXA1 | rs8018823 | 37718177 | 9.70E-09 | rs8018493 | 37644792 | 0.012 | 0.82 | ↑↑ |
| 12 | 18 | LINC01630•DCC•LINC01919 | rs10502966 | 50748499 | 8.40E-10 | rs17410557 | 50776391 | 2.33E-05 | 0.83 | ↑↑ |
| 13 | 19 | BCAM•NECTIN2•TOMM40•APOE•APOC1•APOC1P1 | rs12972970 | 45387596 | 2.50E-09 | rs34342646 | 45388130 | 0.363 | 1 | ↑↑ |

At the pathway level, a total of 21 Gene Ontology (GO) pathways were significantly enriched (FDR 20%) in chronic back pain and 10 pathways were enriched (FDR 20%) in acute back pain in the UK Biobank discovery cohort (Table 2). Importantly, different pathways were identified for acute and chronic pain states. Neurogenesis and synaptic plasticity were significant in chronic back pain while odontogenesis, cardiac muscle depolarization, and immune response through Th2-helpers were significant in acute back pain. Of note, the significance of the odontogenesis pathway is driven by genes linked to connective tissue disorders (*RSPO2*) and bone remodeling (*TNFRSF11B*).^31,32^^33^ Three pathways for chronic pain were replicated (*P* < 0.05) in the HUNT cohort: Spinal cord ventral commissure morphogenesis (GO:0021965), Positive regulation of kinase activity (GO:0033674,) and Positive regulation of transferase activity (GO:0051347). None of the acute pain pathways were replicated in the HUNT cohort.

**Table 2.**
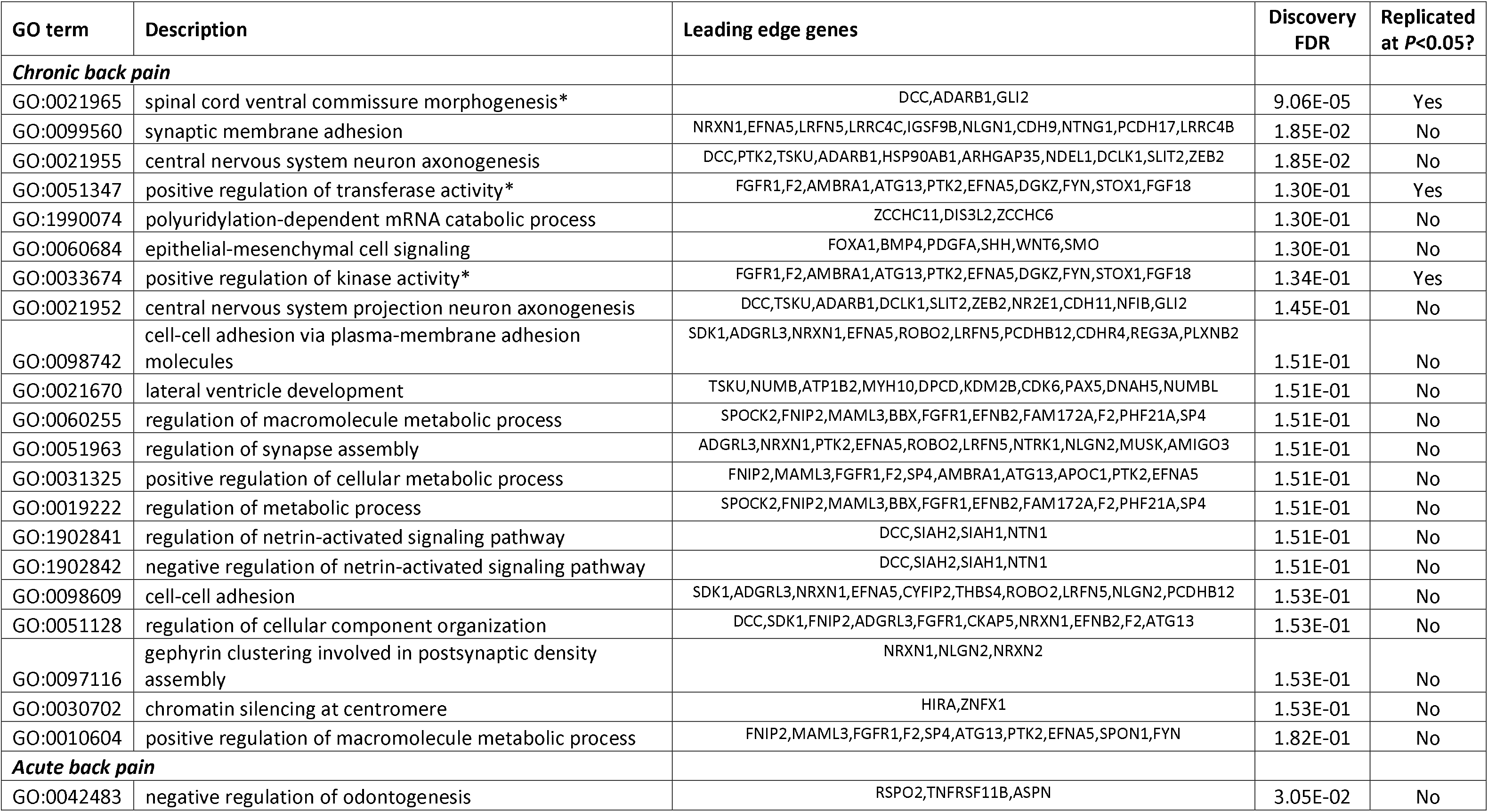
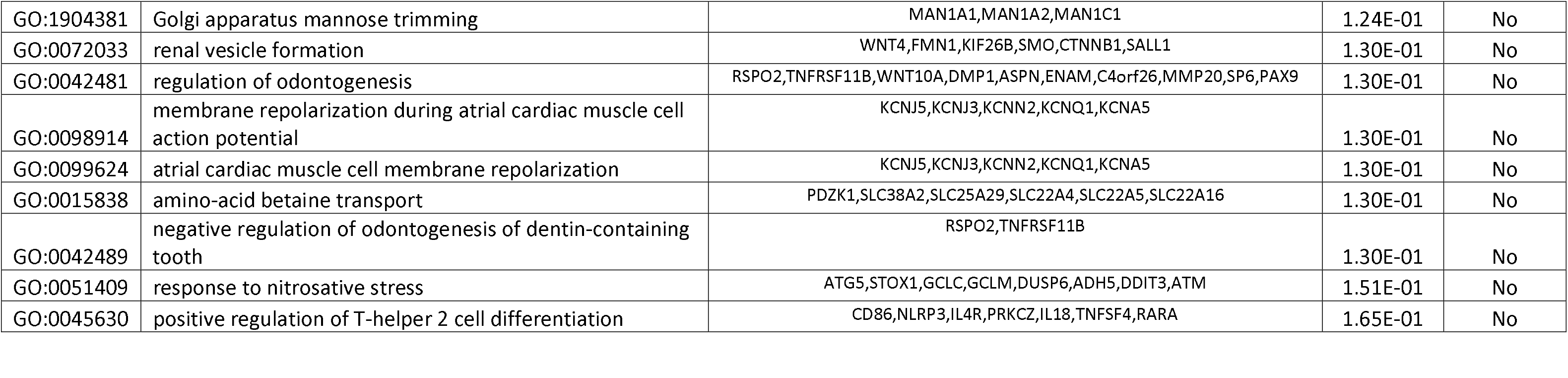
Pathway analysis in the discovery (UK Biobank) and replication (HUNT) cohorts. Pathways that pass the significance threshold FDR 20% in the discovery cohort are shown.

| GO term | Description | Leading edge genes | Discovery FDR | Replicated at $P < 0.05$? |
| --- | --- | --- | --- | --- |
| ***Chronic back pain*** |  |  |  |  |
| GO:0021965 | spinal cord ventral commissure morphogenesis* | DCC, ADARB1, GLI2 | 9.06E-05 | Yes |
| GO:0099560 | synaptic membrane adhesion | NRXN1, EFNA5, LRFN5, LRRC4C, IGSF9B, NLGN1, CDH9, NTNG1, PCDH17, LRRC4B | 1.85E-02 | No |
| GO:0021955 | central nervous system neuron axonogenesis | DCC, PTK2, TSKU, ADARB1, HSP90AB1, ARHGAP35, NDEL1, DCLK1, SLIT2, ZEB2 | 1.85E-02 | No |
| GO:0051347 | positive regulation of transferase activity* | FGFR1, F2, AMBRA1, ATG13, PTK2, EFNA5, DGKZ, FYN, STOX1, FGF18 | 1.30E-01 | Yes |
| GO:1990074 | polyuridylation-dependent mRNA catabolic process | ZCCHC11, DIS3L2, ZCCHC6 | 1.30E-01 | No |
| GO:0060684 | epithelial-mesenchymal cell signaling | FOXA1, BMP4, PDGFA, SHH, WNT6, SMO | 1.30E-01 | No |
| GO:0033674 | positive regulation of kinase activity* | FGFR1, F2, AMBRA1, ATG13, PTK2, EFNA5, DGKZ, FYN, STOX1, FGF18 | 1.34E-01 | Yes |
| GO:0021952 | central nervous system projection neuron axonogenesis | DCC, TSKU, ADARB1, DCLK1, SLIT2, ZEB2, NR2E1, CDH11, NFIB, GLI2 | 1.45E-01 | No |
| GO:0098742 | cell-cell adhesion via plasma-membrane adhesion molecules | SDK1, ADGRL3, NRXN1, EFNA5, ROBO2, LRFN5, PCDHB12, CDHR4, REG3A, PLXNB2 | 1.51E-01 | No |
| GO:0021670 | lateral ventricle development | TSKU, NUMB, ATP1B2, MYH10, DPCD, KDM2B, CDK6, PAX5, DNAH5, NUMBL | 1.51E-01 | No |
| GO:0060255 | regulation of macromolecule metabolic process | SPOCK2, FNIP2, MAML3, BBX, FGFR1, EFNB2, FAM172A, F2, PHF21A, SP4 | 1.51E-01 | No |
| GO:0051963 | regulation of synapse assembly | ADGRL3, NRXN1, PTK2, EFNA5, ROBO2, LRFN5, NTRK1, NLGN2, MUSK, AMIGO3 | 1.51E-01 | No |
| GO:0031325 | positive regulation of cellular metabolic process | FNIP2, MAML3, FGFR1, F2, SP4, AMBRA1, ATG13, APOC1, PTK2, EFNA5 | 1.51E-01 | No |
| GO:0019222 | regulation of metabolic process | SPOCK2, FNIP2, MAML3, BBX, FGFR1, EFNB2, FAM172A, F2, PHF21A, SP4 | 1.51E-01 | No |
| GO:1902841 | regulation of netrin-activated signaling pathway | DCC, SIAH2, SIAH1, NTN1 | 1.51E-01 | No |
| GO:1902842 | negative regulation of netrin-activated signaling pathway | DCC, SIAH2, SIAH1, NTN1 | 1.51E-01 | No |
| GO:0098609 | cell-cell adhesion | SDK1, ADGRL3, NRXN1, EFNA5, CYFIP2, THBS4, ROBO2, LRFN5, NLGN2, PCDHB12 | 1.53E-01 | No |
| GO:0051128 | regulation of cellular component organization | DCC, SDK1, FNIP2, ADGRL3, FGFR1, CKAP5, NRXN1, EFNB2, F2, ATG13 | 1.53E-01 | No |
| GO:0097116 | gephyrin clustering involved in postsynaptic density assembly | NRXN1, NLGN2, NRXN2 | 1.53E-01 | No |
| GO:0030702 | chromatin silencing at centromere | HIRA, ZNFX1 | 1.53E-01 | No |
| GO:0010604 | positive regulation of macromolecule metabolic process | FNIP2, MAML3, FGFR1, F2, SP4, ATG13, PTK2, EFNA5, SPON1, FYN | 1.82E-01 | No |
| ***Acute back pain*** |  |  |  |  |
| GO:0042483 | negative regulation of odontogenesis | RSPO2, TNFRSF11B, ASPN | 3.05E-02 | No |
| GO:1904381 | Golgi apparatus mannose trimming | MAN1A1,MAN1A2,MAN1C1 | 1.24E-01 | No |
| GO:0072033 | renal vesicle formation | WNT4,FMN1,KIF26B,SMO,CTNNB1,SALL1 | 1.30E-01 | No |
| GO:0042481 | regulation of odontogenesis | RSPO2,TNFRSF11B,WNT10A,DMP1,ASPN,ENAM,C4orf26,MMP20,SP6,PAX9 | 1.30E-01 | No |
| GO:0098914 | membrane repolarization during atrial cardiac muscle cell action potential | KCNJ5,KCNJ3,KCNN2,KCNQ1,KCNA5 | 1.30E-01 | No |
| GO:0099624 | atrial cardiac muscle cell membrane repolarization | KCNJ5,KCNJ3,KCNN2,KCNQ1,KCNA5 | 1.30E-01 | No |
| GO:0015838 | amino-acid betaine transport | PDZK1,SLC38A2,SLC25A29,SLC22A4,SLC22A5,SLC22A16 | 1.30E-01 | No |
| GO:0042489 | negative regulation of odontogenesis of dentin-containing tooth | RSPO2,TNFRSF11B | 1.30E-01 | No |
| GO:0051409 | response to nitrosative stress | ATG5,STOX1,GCLC,GCLM,DUSP6,ADH5,DDIT3,ATM | 1.51E-01 | No |
| GO:0045630 | positive regulation of T-helper 2 cell differentiation | CD86,NLRP3,IL4R,PRKCZ,IL18,TNFSF4,RARA | 1.65E-01 | No |

We next investigated how the genetic heritabilities of chronic and acute back pain were distributed across different tissues. We measured the enrichment for the genes corresponding to identified genomic loci expressed in various human tissues. A total of 152 cell types and tissues from Fehtmann *et al*,^34^ grouped into eight categories (adipose, blood and immune, cardiovascular, central nervous system (CNS), digestive, endocrine, musculoskeletal and connective, and other tissues) were included in the analyses (Fig. 2). For chronic back pain, estimates for heritability that exhibited the smallest *P*-values were attributed to genomic regions expressed only in brain regions or brain as a whole (Fig. 2A). Specific parts of the brain that reached the significance threshold (FDR 10%) were the limbic system, parietal lobe, brain stem, cerebral cortex, entorhinal cortex, cerebellum, hippocampus, and metencephalon (Fig. 2B). In contrast, for acute back pain, none of the cell types or tissues reached the statistical significance threshold (Fig. 2C), likely due to low overall heritability of acute back pain. Furthermore, the pattern of tissue-specific partitioned heritabilities for acute back pain appeared to be different from those for chronic back pain, with notable absence of significant signals from brain regions (Fig. 2A and 2C).

**Figure 2.**
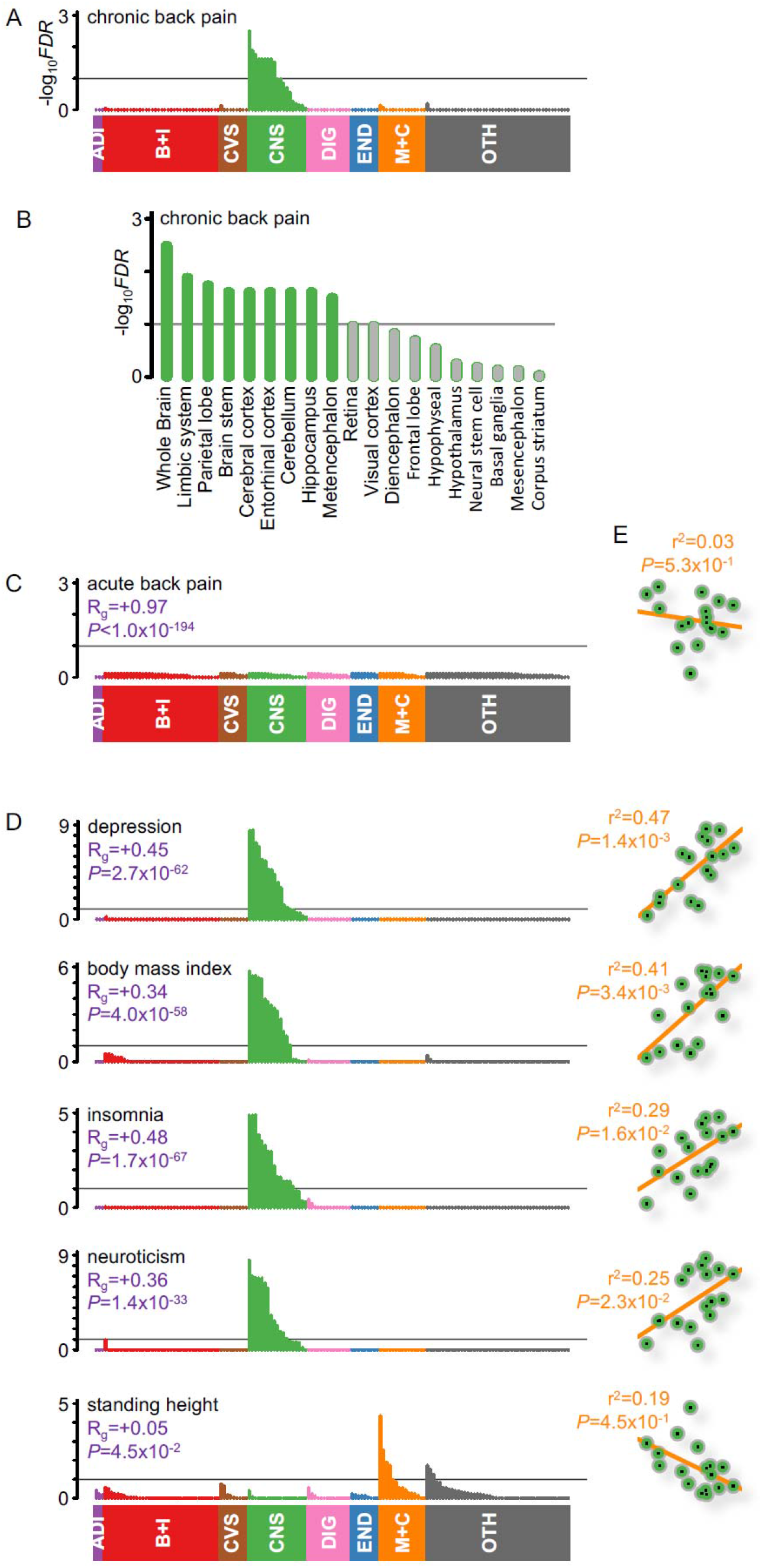
Tissue-specific heritability enrichment for chronic back pain, acute back pain, and other related phenotypes. Genomic tissue-specific annotations for 152 tissues were grouped into eight categories: adipose, ADI (n=3, purple); blood + immune, B+I (n=37, red), cardiovascular, CVS (n=9, brown); central nervous system, CNS (n=19, green); digestive, DIG (n=14, pink); endocrine, END (n=9, blue); musculoskeletal + connective, M+C (n=15, orange); and other, OTH (n=46, grey). Vertical bars in each plot denote –log_10_(FDR) values for enrichment of each tissue. FDR 10% threshold is denoted by a horizontal grey line. **(A, B)** Tissue-specific partitioned heritability for chronic back pain. **(C)** Tissue-specific partitioned heritability for acute back pain, genetic correlation (R_g_) with chronic back pain, and genetic correlation P-value. **(D)** Tissue-specific partitioned heritability for depression, body mass index, insomnia, neuroticism and standing height, genetic correlation (R_g_) with chronic back pain and genetic correlation P-value. **(E)** Brain regions partitioned heritability enrichment correlation between chronic back pain (horizontal axis, not shown) and acute back pain, depression, body mass index, insomnia, neuroticism, and standing height (vertical axis, not shown). Squared correlation coefficient (r^2^) and associated P-value (*P*) are shown in orange.

Next, we used an LD Score regression to estimate genetic correlations between acute and chronic back pain and several selected phenotypes known to be correlated with chronic back pain at the phenotypic level, including body mass index (BMI), insomnia, neuroticism, and depression (Fig. 2D). All of these phenotypes were moderately genetically correlated with chronic back pain, and some of them (BMI, insomnia, and neuroticism) appeared to have significant partitioned heritability in brain tissues, similar to that of chronic back pain (Fig. 2D). In order to look at the similarities/dissimilarities of heritability patterns in brain tissues between chronic back pain and the other phenotypes, we performed a correlation analysis of the heritability estimate coefficients in brain regions (Fig. 2E). A positive correlation would suggest that partitioned heritability is similarly distributed in brain regions between chronic back pain and another phenotype. Interestingly, chronic back pain and acute back pain had strong overall genetic correlation (R_g_=0.97, *P*< 10^−194^), suggesting that genetic heritability for acute pain, although small, largely overlaps with genetic heritability for chronic back pain. However, when considering only brain tissues, no correlation was found between acute and chronic back pain(*P*=0.53, Fig. 2E). Positive correlations of partitioned heritability coefficients in chronic back pain were found for BMI, depression, insomnia, and neuroticism, suggesting that heritability estimates are distributed similarly across brain tissues for these conditions. In contrast, partitioned heritability estimates for brain tissues were not statistically significant in standing height, and did not correlate with heritability estimates for chronic back pain (Fig. 2D–E).

Our findings suggested that chronic back pain is determined by genes predominantly expressed in the brain. To validate our findings, we performed a gene enrichment analysis using data on tissue-specific differential gene expression in mouse assays of inflammatory and neuropathic pain.^35^ Four tissues, namely the whole brain, spinal cord, dorsal root ganglia, and whole blood, were available for investigation in two mouse pain assays: Complete Freund Adjuvant (CFA) for inflammatory pain, and Spared Nerve Injury (SNI) for neuropathic pain. In line with our partitioned heritability findings, we observed enrichment of genes upregulated in the nervous system tissues in both mouse pain models (Fig. 3), suggesting that both neuropathic and inflammatory components contribute to human back pain. Inflammatory pain-related genes that were differentially expressed in the brain, spinal cord, and dorsal root ganglia in mice were significantly enriched in chronic, but not acute, back pain GWAS (Fig. 3A). Similarly, neuropathic pain-related genes that were differentially expressed in the mouse brain were significantly enriched in chronic, but not acute, back pain GWAS (FDR 10%, Fig. 3B). Altogether, heritability of chronic back pain is not only largely mediated by SNPs in genes expressed in the brain, but also by those genes that are differentially-expressed in a pain-conditioned brain.

**Figure 3.**
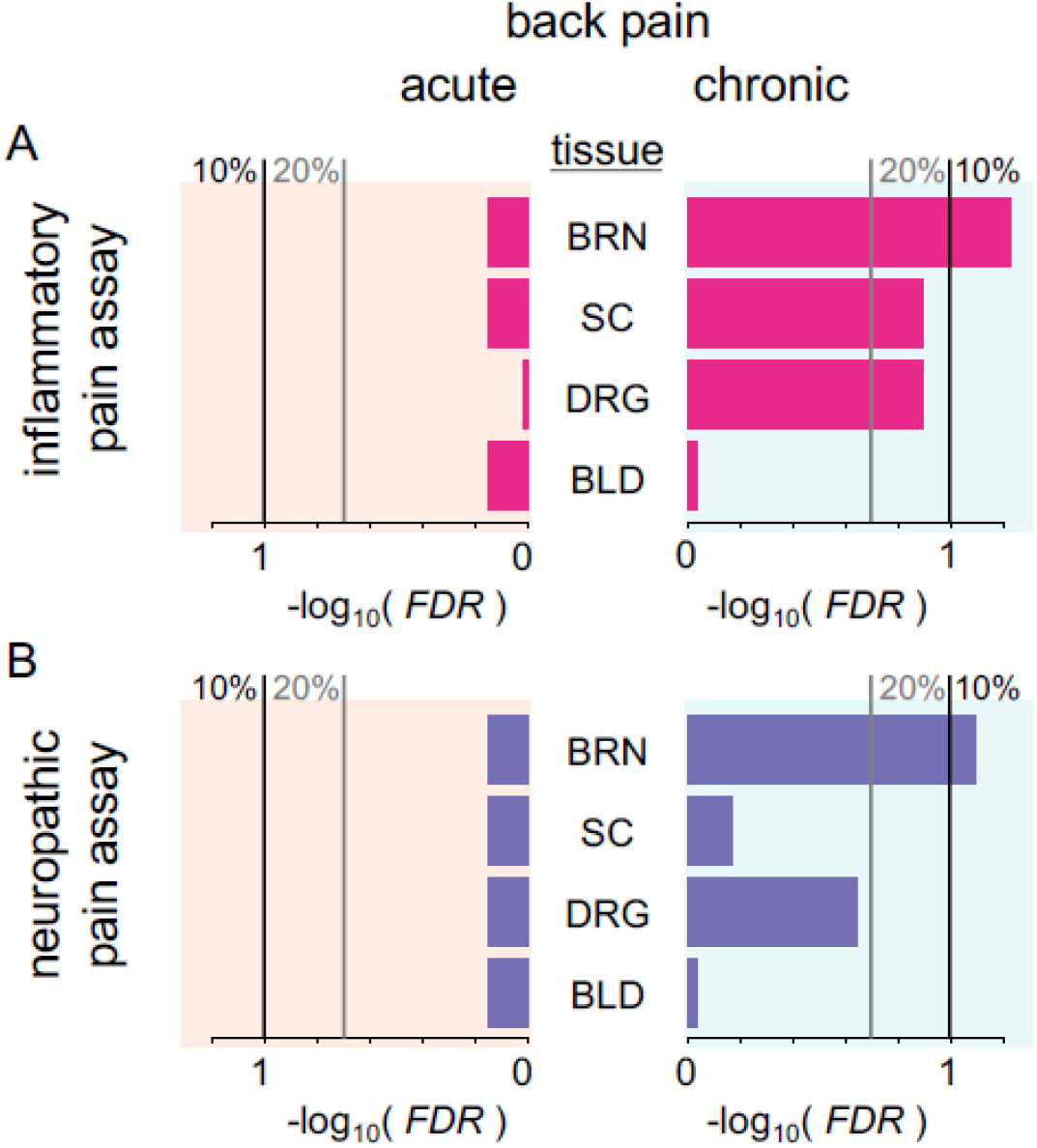
Tissue-specific heritability enrichment for back pain GWAS results using transcriptomic annotations from inflammatory **(A)** and neuropathic **(B)** mouse pain assays. Annotations for genes upregulated in whole brain (BRN), spinal cord (SC), dorsal root ganglia (DRG), and whole blood (BLD) were used to determine heritability enrichment in chronic and acute back pain GWAS. Shown are –log10 FDR-corrected P-values for enrichment of heritability. Vertical black lines indicate FDR 10%; vertical grey lines indicate FDR 20%.

We then hypothesized that epigenetic markers of the 13 genomic loci that reached genome-wide significance (Table 1) in our chronic back pain analyses would be enriched in brain tissues (Fig. 4). To test this hypothesis, we first mapped the significant SNPs onto corresponding genes and defined 13 gene clusters (Fig. 4A). Then we looked at the intersection of SNPs in linkage disequilibrium (LD) with the lead SNP in each cluster with NIH Epigenetics Roadmap activation markers (H3K4me3 for promoter regions,^36,37^ H3K4me1 for enhancers,^36^ and H3K36me3 for transcribed regions^38^) for statistically significant enrichment in 396 different tissues^39^ (Fig. 4B). Our results again aligned with the partitioned heritability findings: 7 out of 13 loci co-localized with epigenetic markers in multiple brain tissues, as evidenced by statistically significant P-values (Bonferroni-corrected *P*<0.05, Fig. 4B). These results suggest that these regions are transcriptionally active in the CNS. Using a different chromatin epigenome mapping (NIH Epigenetic Roadmap chromatin 15-state model),^40^ we found that 9 out of 13 loci had significant enrichment in multiple brain tissues (Fig. 4C). Thus, our epigenetic characterization of the chronic back pain-related variants suggests that expression of the genes in the CNS plays a significant role in the chronic back pain phenotype, followed by genes expressed in the musculoskeletal system (osteoblasts, chondrocytes, and muscles).

**Figure 4.**
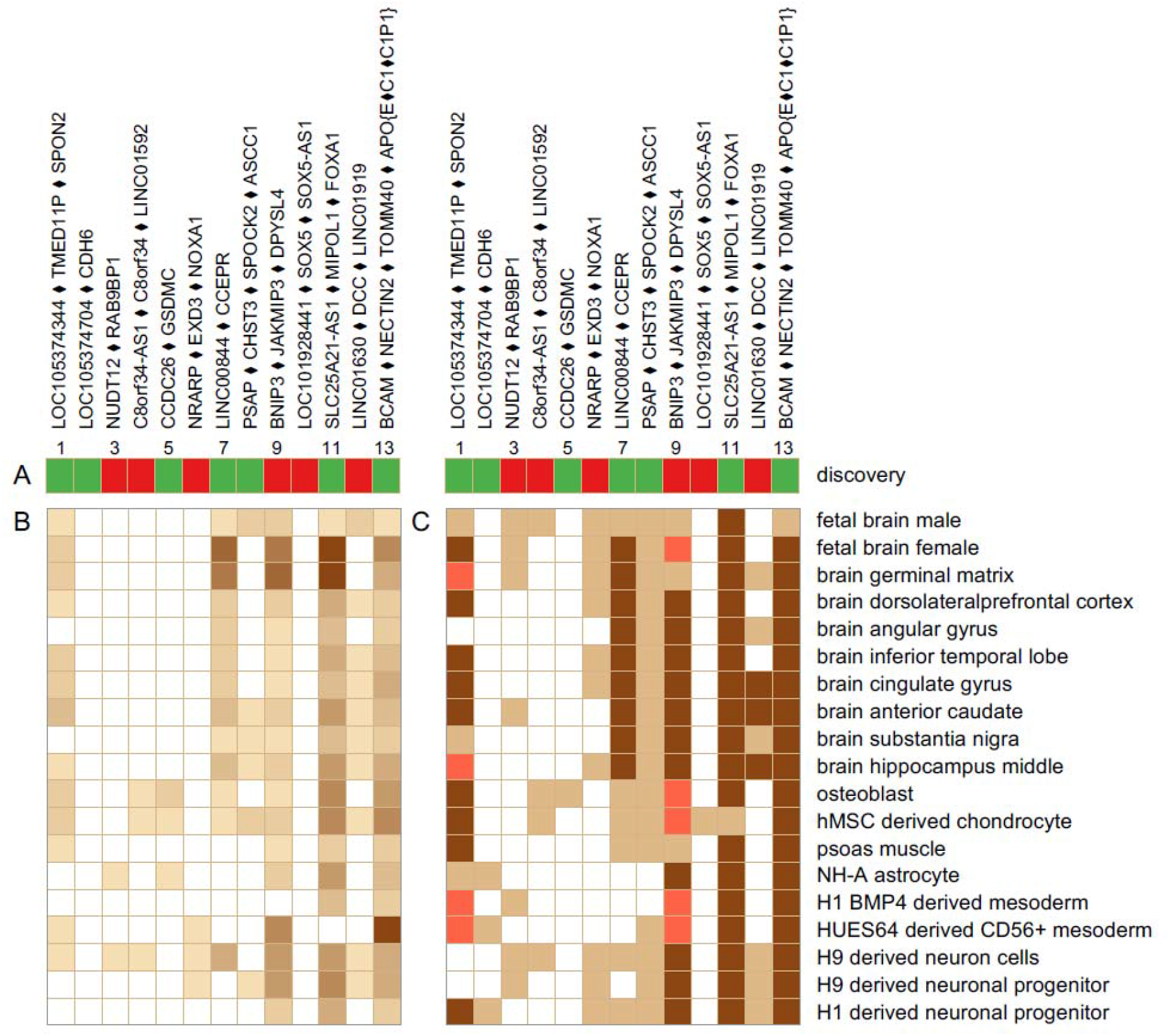
Epigenetic characterization of the GWAS significant 13 chronic back pain loci. **(A)** Gene clusters. Lead SNP effect of minor allele in the discovery cohort: protective (green) or risk (red). **(B)** Intersection of SNPs in LD with lead SNP (r^2^≥ 0.5) with NIH Epigenetics Roadmap activation markers in selected tissues. Markers are: H3K4me3, H3K4me1, H3K36me3. Darker hues for increased signal. **(C)** Intersection of SNPs in LD with lead SNP (r^2^ ≥ 0.5) with NIH Epigenetics Roadmap 15-states chromatin model in selected tissues. States are: transcription start site (state 1; dark brown) and enhancer (states 6 or 7; light brown) indicating active transcription, and bivalent or poised transcription start site (state 10; orange) indicating absence of active ≤ transcription. Statistical significance for ChIP-Seq signal established at *P* 0.05/(3 markers × 13 loci × 19 tissues). Cells are colored white otherwise.

## DISCUSSION

Previous studies suggest substantial differences between acute and chronic back pain in terms of predisposing factors.^41,42^ Our results from two large cohorts indicate the genetic contribution to chronic back pain is greater than to acute back pain, and much of the heritability of chronic back pain can be traced to genes predominantly expressed in the CNS. At the pathway level, we found the enrichment for genes in the spinal cord ventral commissure morphogenesis pathway in both cohorts. We also mapped the genetic component of chronic but not acute pain states to genes differentially expressed in the brain in mouse pain models. Interestingly, we found this enrichment for brain specific differentially-expressed genes in both inflammatory and neuropathic mouse pain models, and also in the spinal cord and in dorsal root ganglia of the inflammatory pain model.

Our observation that acute back pain is significantly less heritable than chronic back pain(0.8% for acute versus 4.6% for chronic, narrow-sense SNP heritability) may be due to a greater role of environmental (e.g. tissue injury) factors in acute pain. In turn, transition to chronic back pain, or development of chronic back pain *per se*, appears to require predisposing genetic and/or epigenetic risk factors. It was observed that epigenetic regulation was still occurring in the forebrains of mice that underwent peripheral nerve injuries long after the injury.^43^

While we found a high genetic correlation between chronic and acute back pain, this observation does not contradict our finding that acute back pain is considerably less heritable than chronic back pain. Indeed, genetic correlation is the genetic covariance, i.e., the covariance between sets of SNP contributions to heritability for two traits divided by the square root of h_1_^2^ and h_2_^2^. This genetic correlation can be large if total heritability of one of the traits (h_1_) due to a set of SNPs is relatively small compared to total heritability of the second trait (h_2_) and when h_1_ constitutes a subset of h_2_ in terms of individual SNP contributions. It is inevitable that some individuals with acute back pain in our study will not resolve their pain over time. It is conceivable that the positive genetic correlation may predispose these individuals to chronic back pain. Genetic correlations between chronic back pain and other phenotypes (BMI, insomnia, neuroticism, and depression) were of moderate strength, and are in line with previous studies’ results showing genetic similarities between these phenotypes.^26,44,45^

The main finding in our study is that heritability of chronic back pain, but not acute back pain, is largely attributed to genes expressed in brain tissues. Involvement of the CNS in chronic back pain is an area of active research,^46^ supported by a significant comorbidity between chronic back pain and psychological disorders.^47^ Indeed, we found genetic overlap of chronic back pain with sleep disorders, neuroticism, and BMI. While this overlap has been previously described in several genetic studies ^22,26,48^), here we show that the overlap is localized in genes predominantly expressed in the CNS. Specifically, we found that chronic back pain heritability is mainly localized in the following brain regions: limbic system, parietal lobe, brain stem, cerebral cortex, entorhinal cortex, cerebellum, hippocampus, and metencephalon. Moreover, the distribution of heritability across brain regions was similar for chronic back pain, insomnia, neuroticism, and BMI, suggesting shared genetic and pathophysiological mechanisms between these phenotypes. Our finding of predominant involvement of brain-expressed genes in chronic back pain, but not acute back pain, corroborates previous studies’ findings suggesting that spontaneous pain in chronic pain patients involves specific spatiotemporal neuronal mechanisms implicating a salient role for the emotional brain, distinct from mechanisms observed for acute experimental pain.^49^ Interestingly, in our study we found only limited evidence of the involvement of musculoskeletal tissues in chronic back pain.

Using the UK Biobank cohort for discovery allowed use of a larger sample size than in previous GWAS studies ^26,27^ and enabled us to identify 13 loci that exceeded a genome-wide significance threshold for chronic back pain. We have replicated seven loci in the HUNT cohort. Of those, 3 loci are novel (Chr5: LOC105374704 • CDH6, Chr5: NUDT12 • RAB9BP1, Chr14: SLC25A21-AS1 • MIPOL1 • FOXA1). Another locus (Chr18: LINC01630 • DCC • LINC01919) has never previously been replicated in an independent cohort.

Our results suggest that chronic pain is much more heritable than acute pain. Furthermore, despite of the strong genetic correlation between acute and chronic pain, the most significant pathways contributing to these pain states seem to be different. Pathway analyses of chronic back pain GWAS revealed enrichment for genes involved in the spinal cord ventral commissure morphogenesis pathway in both the UK Biobank and HUNT cohorts. The ventral white commissure is comprised of A delta and C fibers, which are known to be involved in pain signal transduction. This suggests that molecular mechanisms of nerve fiber growth might be involved in the pathophysiology of chronic back pain.^50^ Interestingly, the most significant pathway in acute back pain, GO:00042483, contains two genes linked to connective tissue disorders (RSPO2) and bone remodeling (TNFRSF11B). ^31,32,33^ It is possible, therefore, that different gene subsets drive the bulk of the onset and persistence of back pain, with the majority of the pathways contributing to acute back pain not necessarily contributing to pain becoming chronic, in addition to a strong environmental component in acute back pain.

A limitation of our study is in the extent to which the phenotypes of interest, acute and chronic back pain, are ascertained in the UK Biobank cohort. Pain intensity, frequency of episodes, and pain medication use were not available for analyses. Additionally, phenotype definitions varied slightly between the UK Biobank and the HUNT cohorts (back pain in the UK Biobank and low back pain in the HUNT). However, we concluded that these differences did not affect the validity of our findings given the high replication success rate of our findings between the two cohorts.

In conclusion, our analyses show that chronic back pain is substantially more heritable than acute back pain, and this heritability is mostly attributed to genes expressed in the brain. Furthermore, our results suggest that different genetic pathways may be responsible for acute and chronic pain manifestation. Molecular pathophysiology of acute back pain is largely contributed to by connective and bone tissue remodeling and inflammatory pathways while chronic back pain is contributed to by neuronal development processes. These results provide substantial insight into physiological systems and pathways responsible for acute and chronic back pain, and should be expanded upon to further to develop targeted treatments.

## METHODS

### The UK Biobank cohort

The UK Biobank cohort was used as a discovery cohort. This cohort is a high-powered prospective study of 500,000 people recruited in the United Kingdom.^51,52^ It is mainly comprised of individuals of Caucasian ancestry whose ages range between 37 and 73 years, with a female to male participant ratio of about 1.2:1. Chronic cases were defined as those answering “yes” to the question “Have you had back pains for more than 3 months?” (field 3571). This question was asked to participants if they answered “yes” for the “Back Pain” option at this question: “In the last month have you experienced any of the following that interfered with your usual activities?” (field 6159). Participants were discarded based on the following criteria: answering “no”, “do not know” or “prefer not to answer” (field 3571), not “White British”, failed genotyping quality control or sex mismatch, and voluntary withdrawal from the study. Acute back pain cases were identified as those answering “no” at field 3571. All other participants (i.e. those who did not met the criteria for acute or chronic back pain) were qualified as controls. Therefore, control individuals might have experienced acute or chronic pain at other bodily sites than in the back. Genotyping, quality control, and genomic imputation in the UK Biobank cohort were previously described.^53^

### The HUNT cohort

The Nord-Trøndelag Health Study (HUNT) cohort was used for replication of results. The HUNT study has been conducted in Nord-Trøndelag County, Norway, in three consecutive waves^54^: HUNT in 1984–1986, HUNT2 in 1995–1997 and HUNT3 in 2006–2008. This work combines baseline data from the second survey, HUNT2,^55^ and the third survey, HUNT3.^54^ All residents of Norway aged 20 years and older were invited to take part in the HUNT2 survey. They were requested to complete a questionnaire on health status and invited to a clinical consultation that included measuring height and weight. In the HUNT3 survey 11 years later, similar information was collected using a questionnaire and a clinical examination. Each participant in the HUNT2 and HUNT3 surveys signed a written informed consent regarding the collection and use of data for research purposes.

One question in the HUNT2 and HUNT3 questionnaires was expressed: ‘During the last year, have you suffered from pain and/or stiffness in your muscles and joints that has lasted for at least 3 consecutive months?’ Each participant answering yes was given the following question: ‘Where did you have these complaints?’ Several body regions were listed. Individuals answering yes to the first question and including the lower back as a relevant region were regarded as having chronic lower back pain.^56^ Acute low back pain cases (n=4,379) were defined as those who had lumbar pain in the last month (at HUNT2), but not had musculoskeletal pain for more than 3 of the past 12 months (at HUNT2). Controls (n=11,309) were defined as those participants without musculoskeletal pain in the last month or musculoskeletal pain for more than 3 of the past 12 months (at HUNT2) and if they did not have musculoskeletal pain for more than 3 of the past 12 months (at HUNT3). Chronic low back pain cases (n=19,760) were defined as those who had lumbar pain for more than 3 of the past 12 months. Controls (n=28,674) had no musculoskeletal pain for more than 3 of the past 12 months at either HUNT2 or HUNT3.

Genotyping of study subjects was performed in three batches using the Illumina HumanCoreExome (Illumina Inc, CA, USA).

## Statistical analyses

Genome-wide association tests in the UK Biobank cohort were performed using a linear mixed model as implemented in BOLT-LMM software, version 2.3.^29,30^ Covariates were sex, age, age^2^, genotyping platform, first 40 genetic principal components, and recruitment centers. Kinship was considered by BOLT-LMM using genotyped position data. Retained SNPs had minor allele frequencies of at least one-in-a-thousand, departed from Hardy-Weinberg equilibrium with *P-*values greater or equal to 10^−12^, and were part of the Haplotype Reference Consortium (HRC) panel.^57^ Association analyses in the HUNT cohort were performed using a mixed logistic regression(SAIGE), adjusted for sex, genotyping batch, and 5 principal components.

Epigenetic data for functional characterization of GWAS significant SNPs was taken from the NIH Roadmap Epigenomics Consortium.^39,40^ At each significant locus, the list of SNPs in high LD (r^2^ ≥ 0.5) with the locus’ lead SNP was retrieved from LDlink^58^ using the Great Britain population as the reference. We used the ‘intersect’ option of the bedtools^59^ to retrieve high-LD SNPs that overlapped with epigenetic features. Two analyses of epigenetic markers were performed: the first one in relation to the overlap of high-LD SNPs with known epigenetic activation marker peaks, and the second one in relation to the overlap with NIH Roadmap’s 15-states chromatin model built from peaks of activation/repression markers, focusing on transcription start sites (state 1), transcription enhancers (states 6 or 7) indicating active transcription, and bivalent or poised transcription start site (state 10) indicating absence of active transcription. For analyses of activation peaks, retained broad peaks had experimental ChIP-Seq evidence P-values ≤ (0.05 / (13×19×3)), correcting for 13 genome-wide significant loci in 19 pain-relevant tissues for 3 epigenetic activation markers (H3K4me3, H3K4me1, and H3K36me3). For chromatin states, we only considered the presence of an overlap with one of these states (since no P-values were available), prioritized as follows: state 1; states 6 and 7; state 10. All data files were downloaded from the following URLs: https://egg2.wustl.edu/roadmap/data/byFileType/peaks/consolidated/gappedPeak/, https://egg2.wustl.edu/roadmap/data/byFileType/chromhmmSegmentations/ChmmModels/coreMarks/jointModel/final/.

Partitioned heritability analyses were performed to determine heritability enrichment in annotated SNP sets using an LD Score Regression (LDSC).^60^ Functional genomic annotations (tissue- and cell-specific gene expression) were retrieved from an online source database comprised of 152 different human tissues.^34^

Additional custom annotations were built from genes overexpressed in two mouse pain assays.^35^ The mouse pain assays were Complete Freund’s Adjuvant, a model for inflammatory pain, and Spared Nerve Injury, a model for neuropathic pain. Quantitation of gene expression from the RNA-Seq data (GEO accession GSE111216) was obtained using STAR^61^ on Genome Reference Consortium’s GRCm38 genome, followed by featureCount^62^ to extract reads per genes. Over-expressed genes in the two pain assays compared to naïve were identified via DESeq2.^63^ A total of three replicate samples per condition (naïve, CFA, SNI) were available. Mouse gene names were translated to human gene names using the BioMart tool.^64^ The top 10% (n=1,663) genes with the greatest positive test statistic (more expression in the pain assay) were retained to build LDSC’s genetic annotation track, as recommended.^60^ The reference or null model annotation track was built from all annotated genes (n=16,629).

For pathway analyses, the summary GWAS data were first analyzed using MAGMA,^65,66^ which aggregated GWAS SNP P-values into gene-level ones, while considering linkage-disequilibrium between the SNPs. MAGMA was also used to deduce pathway-level P-values from gene-level ones. Pathways were sourced from Gene Ontology’s biological processes,^67,68^ obtained from the Bader lab at URL: http://download.baderlab.org/EM_Genesets/December_01_2019/. Pathways in the discovery cohort (the UK Biobank) at the FDR 20% level were ascertained for replication in the replication cohort (HUNT) with P-value of replication smaller or equal to 0.05.

## Data Availability

The datasets used in this work can be downloaded from the following sources.

https://www.ukbiobank.ac.uk/

https://egg2.wustl.edu/roadmap/data/byFileType/peaks/consolidated/gappedPeak/

http://download.baderlab.org/EM_Genesets/December_01_2019/

## Author contributions

LD and AB conceived the paper; AB, MP, and LD designed the analytical plan and experiments. MP, SK, and AEM performed the bioinformatics analyses. AB and MP interpreted the results and wrote the manuscript. AEM, MUL, IH, KH, JAZ, BSW provided the data from the HUNT study. SK, DZ, AEM, MUL, IH, KH, JAZ, BSW, and LD critically reviewed the manuscript. All the authors read and approved the final manuscript.

## Acknowledgments

We would like to thank all participants enrolled in the studies herein. The analysis in the UK Biobank was performed under application 20802. The Nord-Trøndelag Health Study (The HUNT Study) is a collaboration between HUNT Research Centre (Faculty of Medicine and Health Sciences, NTNU, Norwegian University of Science and Technology), Trøndelag County Council, Central Norway Regional Health Authority, and the Norwegian Institute of Public Health. The genotyping was financed by the National Institute of Health (NIH), University of Michigan, The Norwegian Research Council, and Central Norway Regional Health Authority and the Faculty of Medicine and Health Sciences, Norwegian University of Science and Technology (NTNU). The genotype quality control and imputation has been conducted by the K.G. Jebsen Center for Genetic Epidemiology, Department of Public Health andN, Faculty of Medicine and Health Sciences, Norwegian University of Science and Technology (NTNU).

